# The role of institutional trust in preventive and treatment-seeking behaviors during the 2019 novel coronavirus (2019-nCoV) outbreak among residents in Hubei, China

**DOI:** 10.1101/2020.02.15.20023333

**Authors:** Liping Wong, Qunhong Wu, Xi Chen, Zhuo Chen, Haridah Alias, Mingwang Shen, Jingcen Hu, Shiwei Duan, Jinjie Zhang, Liyuan Han

**Affiliations:** Department of Epidemiology, Zhejiang Provincial Key Laboratory of Pathophysiology, School of Medicine, Ningbo University, Ningbo, Zhejiang, China; Centre for Epidemiology and Evidence-Based Practice, Department of Social and Preventive Medicine, Faculty of Medicine, University of Malaya 50603, Kuala Lumpur, Malaysia; Department of Social Medicine, Health Management College, Harbin Medical University, Harbin, Heilongjiang Province, China; Yale School of Public Health, Office: Ste. 301, 60 College St, Mailing Address: PO Box 208034, New Haven, CT 06520; College of Public Health, University of Georgia, Athens, 30602 Georgia, USA; School of Economics, University of Nottingham Ningbo China, 315100 Ningbo, Zhejiang Province, China; Department of Epidemiology and Biostatistics, School of Public Health, Xi’an Jiaotong University Health Science Center, Xi’an, Shanxi, 710061, PR China; College of Food and Pharmaceutical Sciences, Ningbo University, Ningbo 315000, China

**Keywords:** 2019-nCoV, institutional trust, preventive behaviors

## Abstract

**Background:** Since December 2019, pneumonia associated with the 2019 novel coronavirus (2019-nCoV) has emerged in Wuhan, China. The exponential increase of the confirmed number of cases of 2019n-CoV is of great concern to the global community. The fears and panic among residents in the epicenters have prompted diverse responses, which are understudied. During such a crisis, community trust and support for the government and health authorities are important to contain the outbreak. We aimed to investigate the influence of institutional trust on public responses to the 2019-nCoV outbreak.

**Methods:** An anonymous Internet-based, cross-sectional survey was administered on January 29, 2020. The study population comprised all residents currently residing or working in the province of Hubei, where Wuhan is the capital city. The level of trust in information provision and preventive instructions, individual preventive behaviors and treatment-seeking behaviors were queried.

**Findings:** The majority of the participants expressed a great extent of trust in the information and preventive instructions provided by the central government than by the local government. A high uptake of 2019-nCoV preventive measures was found, particularly among people who had been placed under quarantine. Being under quarantine (adjusted odds ratio [OR] = 2.35, 95% confidence interval [CI] 1.80 to 3.08) and having a high institutional trust score (OR = 2.23, 95% CI 1.96 to 2.53) were both strong and significant determinants of higher preventive behavior scores. The majority of study participants (85.7%, n = 3,640) reported that they would seek hospital treatment if they suspected themselves to have been infected with 2019 n-CoV. Few of the participants from Wuhan (16.6%, n = 475) and those participants who were under quarantine (13.8%, n = 550) expressed an unwillingness to seek hospital treatment. Similarly, being under quarantine (OR = 2.36, 95% CI 1.80 to 3.09) and having a high institutional trust score (OR = 2.20, 95% CI 1.96 to 2.49) were two strong significant determinants of hospital treatment-seeking.

**Interpretation:** The results of this study suggest that institutional trust is an important factor influencing adequate preventive behavior and seeking formal medical care during an outbreak. In view of the 2019-nCoV being highly pathogenic and extremely contagious, our findings also underscore the importance of public health intervention to reach individuals with poor adherence to preventive measures and who are reluctant to seek treatment at formal health services.

**Funding:** National Key R&D Program of China, Ningbo Health Branding Subject F und, Sanming Project of Medicine in Shenzhen, K.C. Wong Magna Fund in Ningbo University, National Natural Science Foundation of China, Fundamental Research Funds for the Central Universities, China Postdoctoral Science Foundation, and Natural Science Basic Research Program of Shanxi Province.

**Evidence before this study:** We searched PubMed on January 28, 2020, for articles that describe the trust, preventive practices and health-seeking behaviors related to the 2019 novel coronavirus (2019-nCoV) in China, using the search terms “novel coronavirus,” “institutional trust,” “behavioral change,” “protective behaviors,” and “treatment-seeking” with no language or publication time restrictions. Previously published research discussed the behavioral responses to the severe acute respiratory syndrome (SARS) epidemic and the 2009 pandemic influenza A (H1N1) in Hong Kong. The only report investigating the influence of institutional trust and public responses was published on March 27, 2019, on the Ebola outbreak in the Democratic Republic of the Congo. No previous studies have investigated how trust in the information provision and prevention instructions from the authorities during a disease outbreak has influenced the public’s prevention practices and treatment-seeking in the epicenter of the 2019-nCoV.

## Introduction

Starting on December 8, 2019, several cases of pneumonia were reported in Wuhan, Hubei province, China *(1,2)*. Subsequently, on January 7, 2020 the Chinese Center for Disease Control and Prevention (CDC) identified a novel coronavirus linked to the outbreak, which was subsequently named the 2019 novel coronavirus (2019-nCoV) by the World Health Organization (WHO) *(3)*. On January 23, 2020, the central government of the People’s Republic of China imposed a lockdown in Wuhan in an effort to quarantine the epicenter of the outbreak to prevent an epidemic. Subsequently, two other cities, Enzhou and Chibi, were also locked down. Most recently, on January 29, 2020 the WHO has declared the 2019-nCoV a global health emergency.

In this ongoing severe case of escalating infections and deaths, healthcare providers in China are working around the clock treating patients and preventing casualties, while scientists in China and around the globe are racing to find out more about the coronavirus to prevent it becoming a worldwide pandemic. Studies published in the past weeks have led to a better worldwide understanding of the epidemiology, clinical and genomic characteristics of the 2019-nCoV *(4,5)*. However, counteracting the newly discovered 2019-nCoV will entail multifaceted control strategies. Global outbreaks like the severe acute respiratory syndrome (SARS) epidemic and the 2009 pandemic influenza A (H1N1) have brought attention to the importance of understanding community responses in outbreak control *(6)*.Understanding local community responses is vital to provide insights into the development of risk communication messages to the general public for outbreak prevention and control *(6,7)*.Therefore, investigating the public response to the outbreak is as equally important as epidemiological, clinical, and genomic research.

The lesson learned from the recent 2018-2019 Ebola outbreak suggests that institutional trust is of central importance in effective public health intervention *(8)*. Lack of institutional trust may lead to refusal to comply with a preventive or curative intervention which may result in an increased risk of both acquiring and spreading the disease *(8)*. As noted in the statement from the second meeting of the Emergency Committee regarding the outbreak of 2019-nCoV on January 30, the implementation of comprehensive risk communication strategies to enhance public health measures for containment of the outbreak is vital *(9)*.Therefore, it is of utmost importance that people in the epicenter, particularly in the quarantine area, have full trust in the government institutions to enable successful delivery of risk communication and practice of health protection behaviors.

Currently, over a month since the onset of the 2019-nCoV, the epidemic is still spreading rapidly with escalating confirmed cases and deaths. In consideration of the importance of understanding the community responses, this study aimed to investigate the role of institutional trust in individual preventive and treatment-seeking behaviors during the 2019-nCoV outbreak, which to date has claimed over 1000 lives and has infected over 60,000 people in China.

## Methods

### Study design and participants

An anonymous Internet-based, cross-sectional survey commenced on January 29, 2020. The study population comprised adults ≥18 years of age and residing or working in the province of Hubei, where Wuhan is the capital city. This cross-sectional study was performed in accordance with the STROBE statement.

### Procedures

A snowballing sampling technique was used to recruit the participants. The weblink to the survey was first circulated to the academic staff and students at the Ningbo Medical University, who are from diverse geographical locations of origin. They were instructed to employ their social networks to circulate the link to people residing or working in the province of Hubei. Upon completing the survey, a note to encourage participants to disseminate the survey link to all known contacts in Hubei province was included. Participants were remunerated upon completion of the survey.

The questionnaire was developed in English and translated into Chinese. Independent experts reviewed and validated the translation. The questionnaire was also face validated by local experts and pilot tested. The survey consisted of sections that assessed demographic background, institutional trust, and 2019-nCoV-related preventive and health-seeking behaviors.

Institutional trust was assessed using a series of questions (eight items) that assessed participants’ trust in 2019-nCoV-related information (i.e. the reported number of confirmed cases, deaths, etc.) and preventive instructions (i.e. self-quarantine, provincial quarantine, extended time off, etc.) given by the local, provincial, and central governments. They were also queried about their trust in the information provision and the preventive instructions from healthcare providers. The response options were recorded on a 4-point Likert scale with items scored as either 0 (not at all), 1 (a small extent), 2 (moderate), or 3 (a great extent). The possible score ranged from 0 to 24, with higher scores indicating higher levels of trust.

The section on Preventive behaviors consisted of five parts (14-items), namely: (1) direct avoidance, (2) social interaction avoidance, (3) physical contact avoidance, (4) public space avoidance, and (5) personal protection. The response options were recorded on a 3-point Likert scale with items scored as either 0 (never), 1 (occasionally), or 2 (frequently). The possible score ranged from 0 to 28, with higher scores indicating higher levels of preventive behaviors.

The section on health-seeking behaviors queried participants about the health-seeking behaviors that they would adopt if they suspected themselves to have been infected with 2019-n CoV. The health-seeking behaviors queried were 1) seeking treatment in the hospital, 2) traditional healing, and 3) self-healing (i.e. exercise, high dose of vitamins, plenty of fresh fruit and vegetables). The answers included two options, “yes” and “no”.

### Statistical analyses

Normality testing was performed using the Kolmogorov-Smirnov test. The scores of institutional trust and preventive behaviors were not normally distributed, therefore all results were expressed as the median and interquartile range (IQR). Other descriptive statistics, such as frequency tables, charts, and proportions were used for data summarization. The reliability of the institutional trust and preventive behavior items was evaluated by assessing the internal consistency of the items representing the scores. The 8-item institutional trust and the 14-item preventive behaviors had a reliability (Cronbach’s α) of 0.944 and 0.653, respectively. Multivariable logistic regression was used to determine factors influencing preventive behavior scores and conventional treatment-seeking. Variables that were significant on a chi-square (χ^2^) test were selected for multivariable logistic regression analysis and included in the model using a simultaneous forced-entry method. Odds ratios (OR), 95% confidence intervals (95% CI), and *p* values were calculated for each independent variable. The model fit was assessed using the Hosmer–Lemeshow goodness-of-fit test.^10^ All statistical analyses were performed using the Statistical Package for the Social Sciences, version 20.0 (IBM Corp., Armonk, NY, USA). A *p*-value of less than 0·05 was considered statistically significant.

### Ethics Approval

This survey was part of a continuing public health outbreak investigation and thus considered exempt from institutional review board approval.

## Results

The survey link was disseminated on January 29, 2020, and by January 30 a total of 4,393 responses were received. The final number of responses included in data analyses was 4,245, after data cleaning to remove invalid responses. The demographic characteristics of the participants are shown in the first column of Table 1. Most of the participants were from the city of Wuhan (67.3%) and most (93.6%) reported that they were currently under quarantine.

**Table 1.**
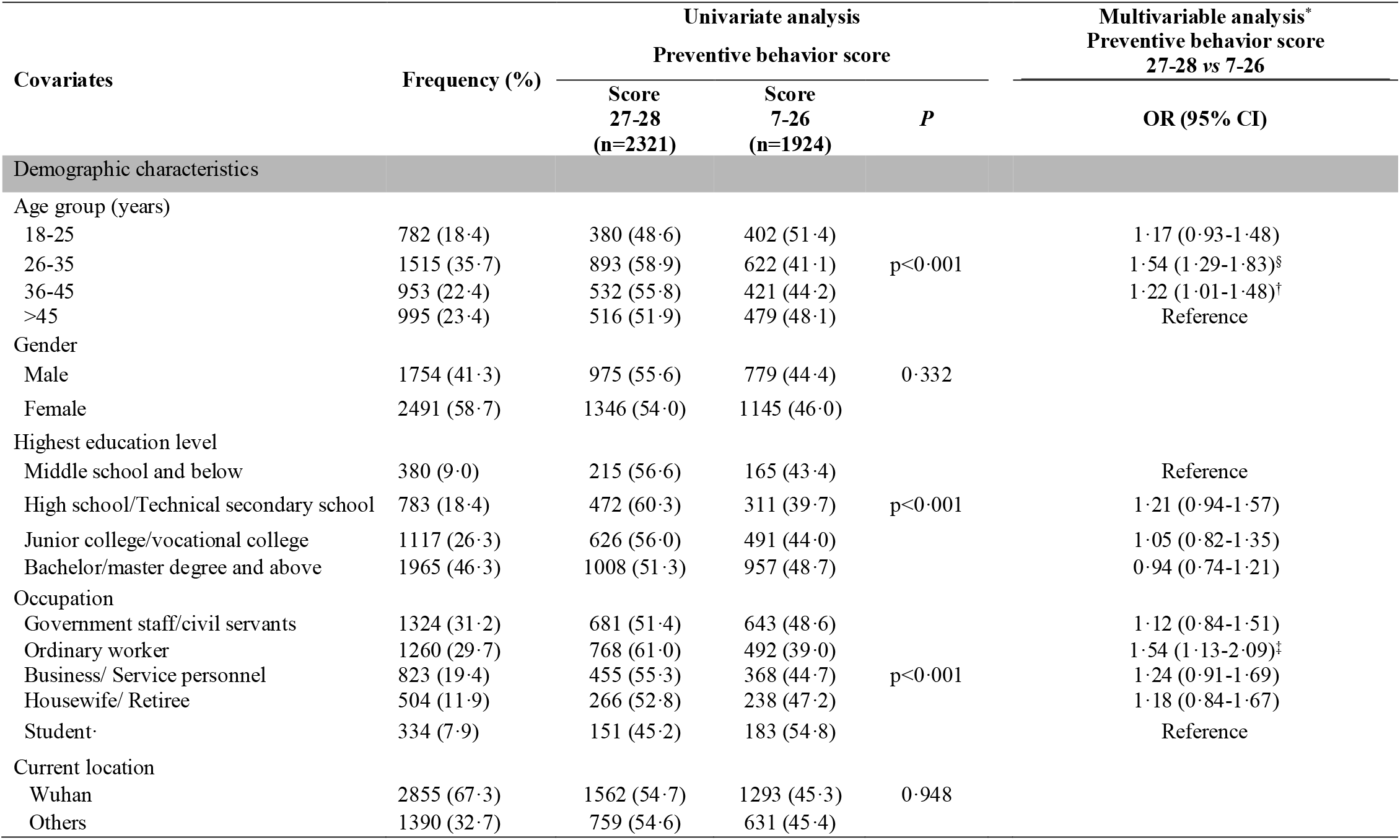
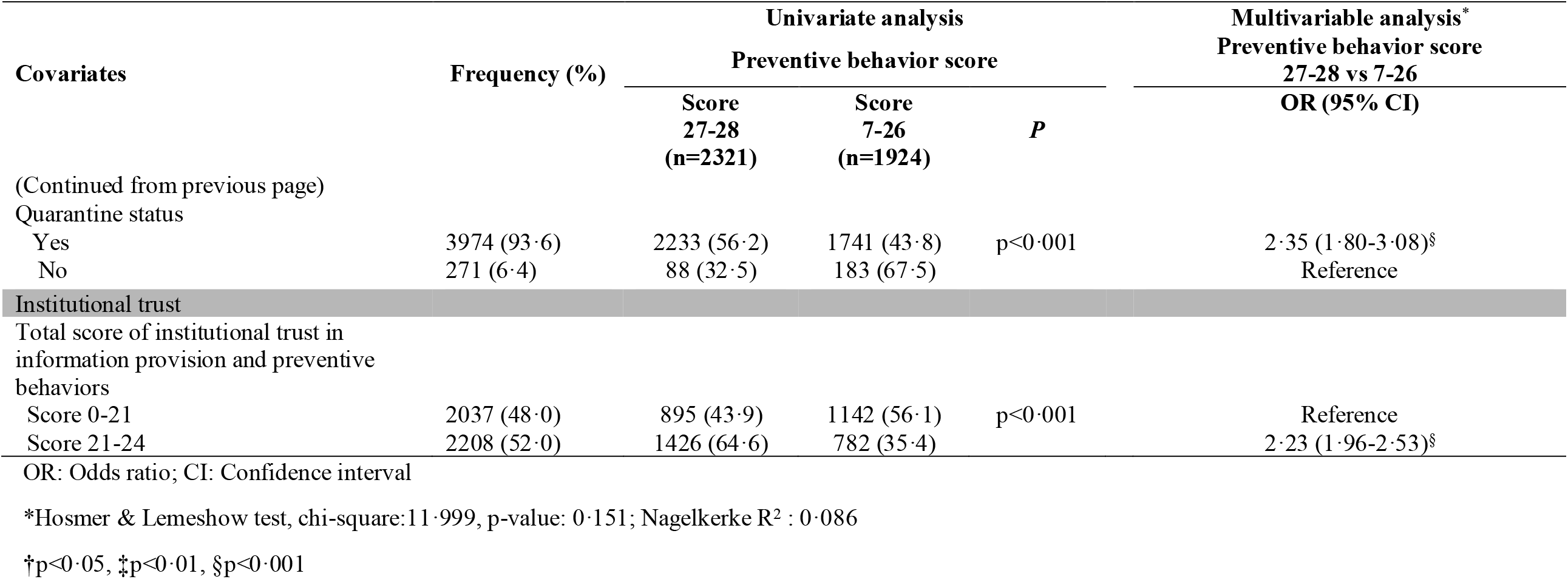
Demographic characteristics and factors associated with preventive behaviors (N=4245)

Figure 1 shows the results of institutional trust for both information provision and preventive instruction. The majority of the participants expressed a great extent of trust in the information provision (72.4%) and preventive instructions (78.5%) from the central government authority. The level of trust in the information provision (68.4%) and preventive instruction (72.1%) from healthcare providers were lower than that of the central government authority. An even lower proportion expressed a great extent of trust in information provision (48.0%) and preventive instructions (59.7%) given by the local authority. The total scores of institutional trust for information provision and preventive instruction ranged from 0 to 24. The median score was 22 (IQR 18 to 24). The total scores was categorized into two groups, namely 22-24 or 0-21, based on the median split; as such, a total of 2,208 (52.0%) were categorized as having a score of 22-24 and 2,037 (48.0%) had a score of 0-21.

**Figure 1.**
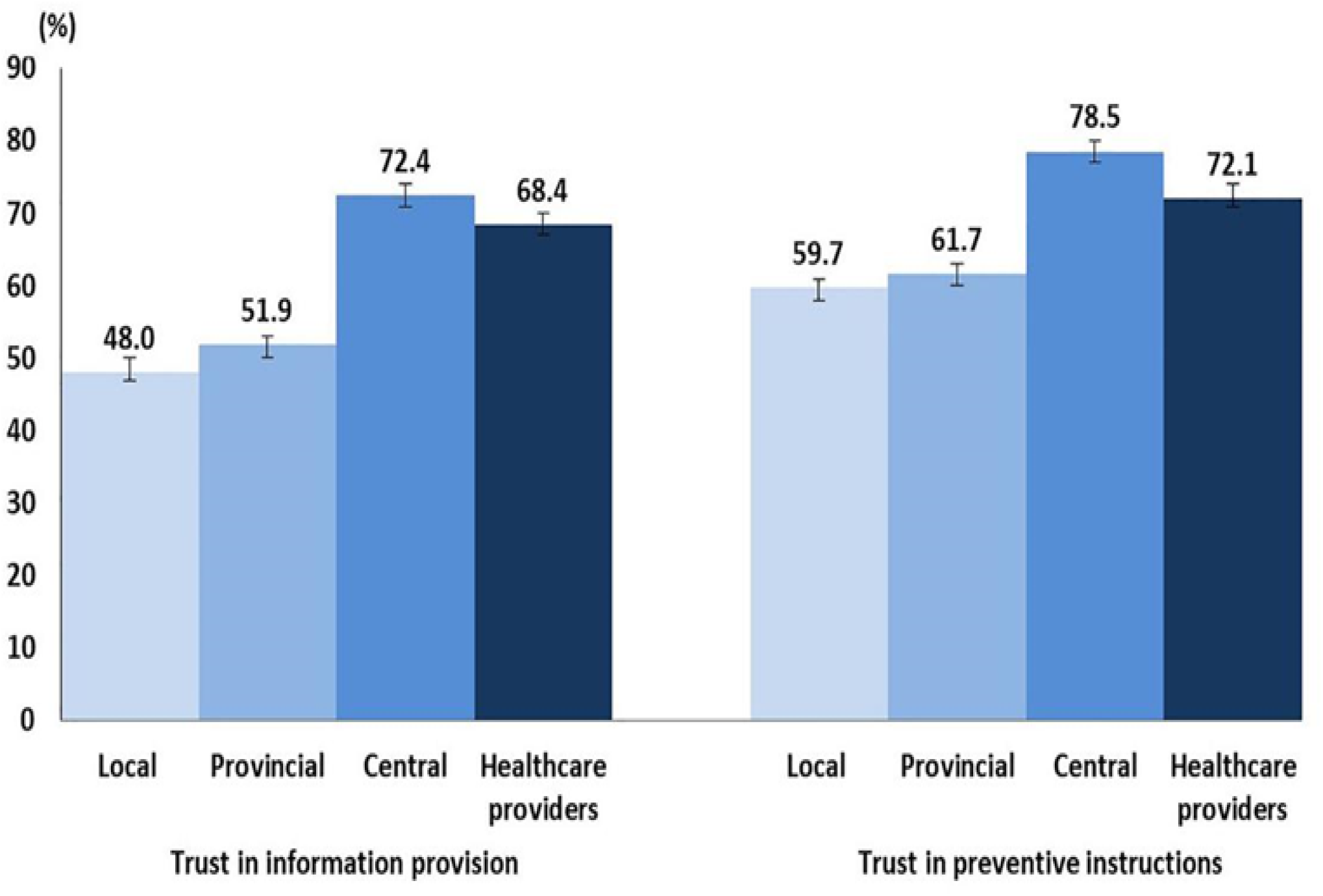
Institutional trust for information provision and preventive instruction (N=4245)

Figure 2 shows the distribution of self-reported preventive behaviors. The uptake of preventive behaviors was high in the study population. However, only 40% reported wearing a face mask at home when they were with other household members. The scores of preventive behaviors in the study population ranged from 7 to 28. The median score was 27 (IQR 26-28). The preventive behaviors scores were categorized as a score of 27 to 28 or 7 to 26 based on the median split; as such, a total of 2321 (54·7%) were categorized as having a score of 27 to 28 and 1924 (45·3%) had a score of 7 to 26. Table 1 shows the univariate and multivariable analyses of factors associated with preventive behavior scores. In the univariate analysis, there were significant associations between the preventive behavior scores and age group, highest educational attainment, occupational types, quarantine status, and the total score of institutional trust. In the multivariate analysis, under quarantine (adjusted odds ratio [OR] = 2·35, 95% confidence interval [CI] 1·80 - 3·08) and having high institutional trust score (OR=2·23, 95% CI 1·96 −2·53) were two strong significant determinants of higher preventive behavior scores. Participants of age group 26-35 years (OR=1·54, 95% CI 1·29 −1·83) and 36 -45 years (OR=1·22, 95% CI 1·01 - 1·48) had significantly higher preventive behaviour scores than those >45 years old.

**Figure 2.**
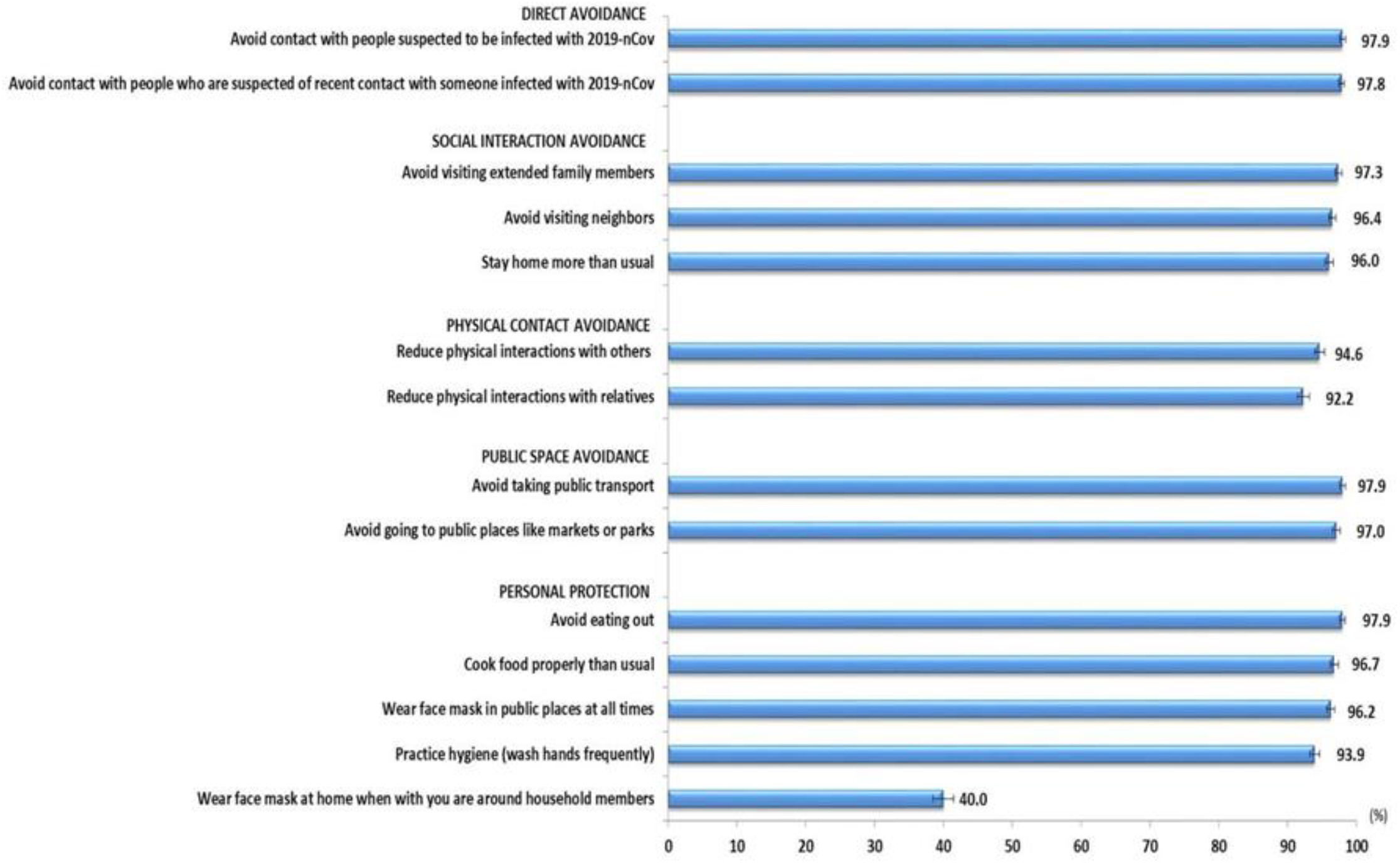
Proportion of Frequent responses for preventive behaviours (N=4245)

Figure 3 shows the treatment-seeking behaviours of the study population. Most participants (85.7%, n = 3,640) reported that they would seek treatment at the hospital, whereas 42% (n = 1,782) reported that they would seek traditional healing, and 62.4% (n = 2,651) reported a preference for self-treatment. A total of 462 (46.3%) participants aged >45 years reported a preference for using traditional healing (χ^*2*^ = 13.649, degrees of freedom = 3, p = 0.011). There was no significant difference in the use of self-healing by participants’ age. Table 2 shows the univariate and multivariable analyses of factors associated with treatment-seeking in the hospital. In the univariate analysis all of the factors studied, except for the age group, were significantly associated with treatment-seeking in the hospital. Of note, 16.6% (n = 475) of the participants from the city of Wuhan reported no intention of seeking treatment in the hospital, whereas 13.8% (n =550) of participants who were in the quarantine area reported no intention of seeking treatment in the hospital. In the multivariable analysis, similarly, being quarantined (OR = 2.36, 95% CI 1.80–3.09) and having a high institutional trust score (OR = 2.20, 95% CI 1.96–2.49) were two strong significant determinants of treatment-seeking in the hospital.

**Table 2.** Factors associated with treatment-seeking in the hospital (N=4245)

| Covariates | Frequency (%) | Univariate analysis |  |  | Multivariable analysis* |
| --- | --- | --- | --- | --- | --- |
|  |  | Treatment-seeking in hospital |  |  | Treatment seeking in hospital<br>Yes vs No |
|  |  | Yes<br>(n=3640) | No<br>(n=605) | P | OR (95% CI) |
| Demographic characteristics |  |  |  |  |  |
| Age group (years) |  |  |  |  |  |
| 18-25 | 782 (18.4) | 655 (83.8) | 127 (16.2) | 0.271 |  |
| 26-35 | 1515 (35.7) | 1297 (85.6) | 218 (14.4) |  |  |
| 36-45 | 953 (22.4) | 824 (86.5) | 129 (13.5) |  |  |
| >45 | 995 (23.4) | 864 (86.8) | 131 (13.2) |  |  |
| Gender |  |  |  |  |  |
| Male | 1754 (41.3) | 1549 (88.3) | 205 (11.7) | p<0.001 | 1.01 (0.89-1.15) |
| Female | 2491 (58.7) | 2091 (83.9) | 400 (16.1) |  | Reference |
| Highest education level |  |  |  |  |  |
| Middle school and below | 380 (9.0) | 340 (89.5) | 40 (10.5) | p<0.001 | Reference |
| High school/Technical secondary school | 783 (18.4) | 709 (90.5) | 74 (9.5) |  | 1.22 (0.94-1.58) |
| Junior college/vocational college | 1117 (26.3) | 977 (87.5) | 140 (12.5) |  | 1.12 (0.87-1.43) |
| Bachelor/master degree and above | 1965 (46.3) | 1614 (82.1) | 351 (17.9) |  | 1.04 (0.81-1.32) |
| Occupation |  |  |  |  |  |
| Government staff/civil servants | 1324 (31.2) | 1105 (83.5) | 219 (16.5) | 0.002 | 1.18 (0.92-1.52) |
| Ordinary worker | 1260 (29.7) | 1110 (88.1) | 150 (11.9) |  | 1.66 (1.28-2.16)<sup>‡</sup> |
| Business/ Service personnel | 823 (19.4) | 718 (87.2) | 105 (12.8) |  | 1.38 (1.06-1.81)<sup>†</sup> |
| Housewife/ Retiree | 504 (11.9) | 434 (86.1) | 70 (13.9) |  | 1.19 (0.88-1.61) |
| Student | 334 (7.9) | 273 (81.7) | 61 (18.3) |  | Reference |
| Current location |  |  |  |  |  |
| Wuhan | 2855 (67.3) | 2380 (83.4) | 475 (16.6) | p<0.001 | Reference |
| Others | 1390 (32.7) | 1260 (90.6) | 130 (9.4) |  | 0.94 (0.82-1.08) |
| Quarantine status |  |  |  |  |  |

(Table 2 continues on next page)
| Covariates | Frequency (%) | Univariate analysis |  |  | Multivariable analysis* |
| --- | --- | --- | --- | --- | --- |
|  |  | Treatment-seeking in hospital |  |  | Treatment seeking in hospital |
|  |  | Yes<br>(n=3640) | No<br>(n=605) | *P* | Yes *vs* No<br>OR (95% CI) |
| (Continued from previous page) |  |  |  |  |  |
| Yes | 3974 (93.6) | 3424 (86.2) | 550 (13.8) | 0.005 | 2.36 (1.80-3.09)‡ |
| No | 271 (6.4) | 216 (79.7) | 55 (20.3) |  | Reference |
| Institutional trust |  |  |  |  |  |
| Total score of institutional trust in information provision and preventive behaviors |  |  |  |  |  |
| Score 0-21 | 2037 (48.0) | 1604 (78.7) | 433 (21.3) | p<0.001 | Reference |
| Score 21-24 | 2208 (52.0) | 2036 (92.2) | 172 (7.8) |  | 2.20 (1.94-2.49)‡ |

OR: Odds ratio; CI: Confidence interval
\*Hosmer & Lemeshow test, chi-square:2.231, p-value: 0.973; Nagelkerke R<sup>2</sup> : 0.079
†p<0.05, ‡p<0.001

**Figure 3.**
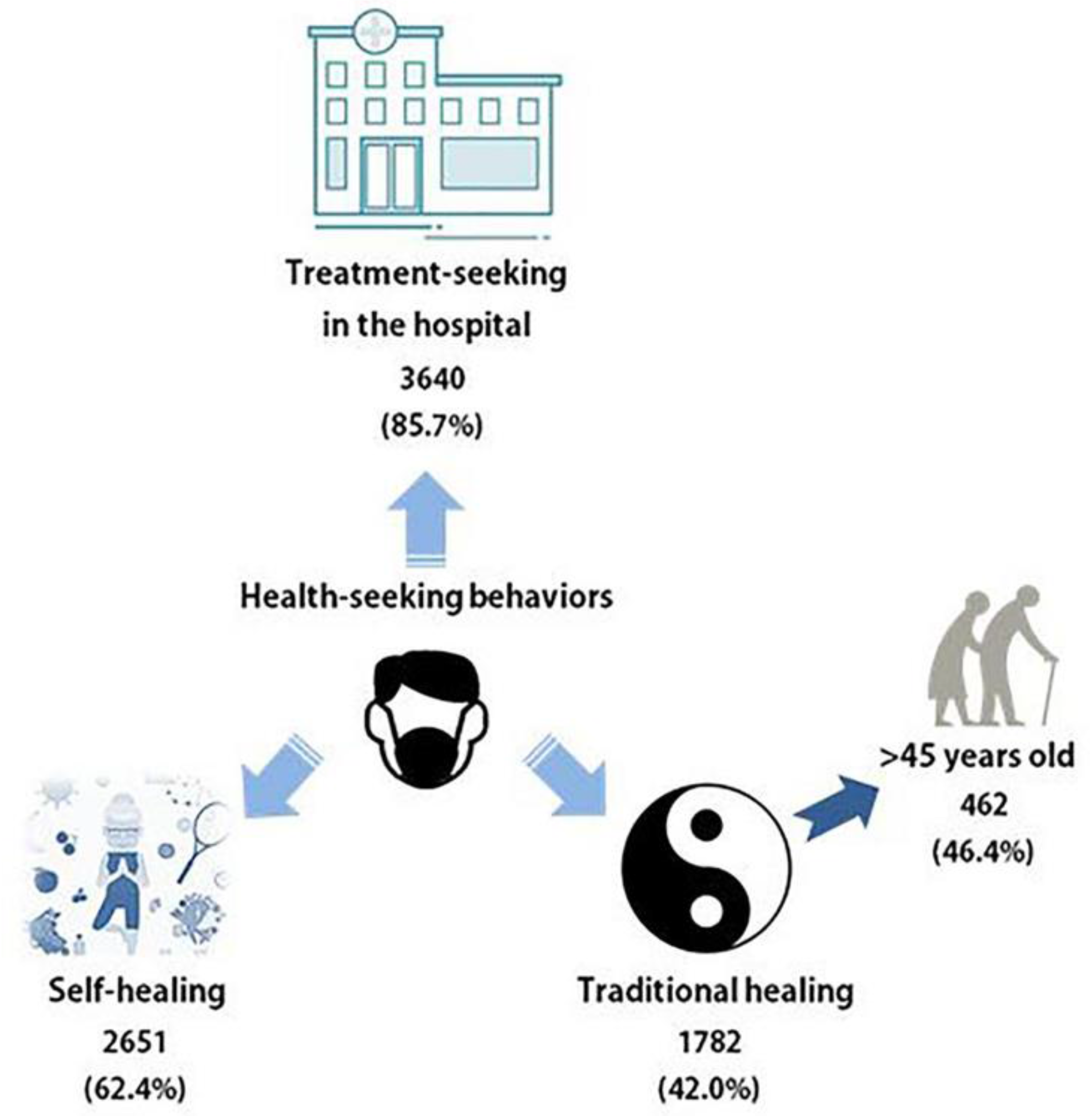
Treatment-seeking behaviors (N=4245)

## Discussion

With the aim of understanding the public behaviour in prevention measures in this crisis that has now become a worldwide concern, this study collected data at the 2019n-CoV outbreak epicentres during a period of exponential growth of the epidemic. This study found a considerably high level of institutional trust among the public in the epicenters. However, trust in the local authority was lower than that in the provincial and central government. This finding is in contrast to the study on the Ebola outbreak in Congo, where their public expressed higher trust in their local authorities (8). There is a need to enhance trust in the local authority, as they are the frontline in service delivery during the outbreak. Lack of trust in the local authority results in poor cooperation, thus undermining contact tracing and adherence to recommended public health interventions *(11,12)*. Healthcare providers can be strong advocates for outbreak prevention efforts in these communities, considering that many expressed a high level of trust in them.

On a positive note, this study revealed that the people in the epicenters adopted a high level of prevention measures. As noted, however, a relatively small proportion reported using face masks at home when they were around household members. Wearing a face mask in public places and at home when around other household members is advisable based on CDC recommendations *(13)*. In consideration that human-to-human transmission among close family members has been reported in overseas countries such as Germany, Japan, and Vietnam, the community in Hubei province and the epicenters should be made aware of the essential importance of wearing face masks when they are around other household members.

The main strength of this study is the finding of the strong influence of institutional trust on overall preventive behaviors, in good agreement with previous studies *(8,11)*. As the 2019-nCoV epidemic is still growing exponentially, continued strengthening of institutional trust, and in particular, increasing the trust in the local authority, is essential for emergency management. outbreak control.

The study also found that uptake of prevention practices was poorer among older members of the public, which warrants serious attention. It has been found that older adults affected by 2019-nCoV are likely to have a higher risk of complications and mortality *(4)*. There is a need to find out whether a lack of appropriate prevention practices among people of older age is due to knowledge deficiency or to their being out-of-reach of current public health intervention.

The finding that nearly 15% of people would be reluctant to seek treatment in the hospital if they were suspected to have been infected with 2019-nCoV is clinically important and worrisome. Of utmost importance, a proportion of those participants are in the city of Wuhan and currently under quarantine. The preference for using traditional healing among a minority of the older respondents is also of concern. Refusal to seek hospital treatment not only leads to progress to serious respiratory distress and can be life-threatening, but also increases the chances of spreading the disease. People who favor traditional healing should be made aware that seeking conventional treatment in the hospital and obeying the quarantine order are the most appropriate actions in containing this outbreak. Finally, the finding of the strong influence of institutional trust on seeking conventional treatment again amplifies the immense importance of building trust between the public and the government authorities.

This current study has several limitations that should be considered. The first pertains to the cross-sectional nature of the study. Thus, it cannot be used to infer causality. Second, the responses were based on self-report and may be subject to self-reporting bias and a tendency to report socially desirable responses. Therefore, the results should be interpreted with caution. Third, the snowballing method used in this study can lead to selection bias. Nevertheless, the demographic distributions of our study population resembles of the general population in Hubei province. Despite these limitations, the study data contributes tremendously to understanding of public responses, especially now that the epidemic is growing exponentially.

In conclusion, bridging the trust gaps between the public and local authorities in the epicentres is crucial. It is of utmost urgency to carry out public health interventions to reach out to individuals with poor adherence to preventive measures and who are reluctant to seek conventional medical care. Considering the extremely contagious nature of the 2019-nCoV, a slight incompliance by even a small portion of the population may have grave consequences and contribute to the continued exponential increase of the outbreak cases.

## Data Availability

The data used to support the findings of this study are available from the corresponding author upon request.

## Acknowledgments

We thank the study participants for their prompt responses. Special acknowledgment for participants who have helped to share the survey link to others in their contact lists.

## Author contributions

QW, XC, ZC, HA, MS, JH, SD and JZ conceived the study and designed the study in collaboration. LPW and LH analyzed the data and wrote the first draft of the manuscript. All authors interpreted the data and contributed to subsequent drafts of the manuscript, and all authors have seen and approved the final version.

## About the Author

Dr. Wong is an epidemiologist at the Department of Social and Preventive Medicine, Faculty of Medicine, University of Malaya, Malaysia. Her primary research has centred almost exclusively on preventive and behavioral medicine, and social epidemiology. She has a great deal of interest in public responses to emerging infectious diseases.

